# Psychotic symptoms are associated with elevated tau PET signal in the amygdala independent of Alzheimer’s disease clinical severity and amyloid burden

**DOI:** 10.1101/2024.01.12.24301221

**Authors:** Aubrey S. Johnson, Galen Ziaggi, Anna C. Smith, Hannah Houlihan, Lauren B. Heuer, Diana S. Guzmán, Amarachukwu Okafor, Edward D. Huey, Daniel Talmasov, Frank Provenzano, William C. Kreisl, Patrick J. Lao, the Alzheimer’s Disease Neuroimaging Initiative

**Affiliations:** Taub Institute for Research on Alzheimer’s disease and the Aging Brain, Gertrude H. Sergievsky Center, Department of Neurology, Columbia University Irving Medical Center, New York, NY 10032; Department of Psychiatry and Human Behavior, Alpert Medical School of Brown University, Providence, RI 02906; Departments of Neurology and Psychiatry, Columbia University Irving Medical Center, New York, NY 10032

## Abstract

**Background:** Psychosis in Alzheimer’s disease (AD) is associated with worse outcomes, yet no established biomarkers exist for early diagnosis and intervention. We compared tau PET burden across older individuals with and without psychotic symptoms.

**Methods:** [^18^F]AV1451 tau PET binding was compared between 26 Alzheimer’s Disease Neuroimaging Initiative (ADNI) subjects with psychotic symptoms (delusions and/or hallucinations) and 26 ADNI subjects without psychotic symptoms, matched for age, sex, race/ethnicity, and clinical severity. Tau was assessed on a region-of-interest and voxel level, corrected for amyloid PET burden.

**Results:** Tau was greater in individuals with psychotic symptoms in the amygdala in region-of-interest analyses, and in amygdala, thalamus, putamen, right hippocampus, right entorhinal cortex, and right frontal cortex in voxel-based analyses. When considering different onset and type of psychotic symptoms, tau binding was greatest in those with concurrent delusions.

**Conclusion:** Elevated tau in limbic regions may be relevant for psychotic symptoms in aging and AD.

## Introduction

Psychosis is a symptom cluster including false beliefs (e.g., delusions) and/or a false perception of events or objects in the surrounding environment (e.g., hallucinations) [1]. The prevalence of psychotic symptoms within Alzheimer’s Disease (AD) is between 40-60%, typically developing in advanced clinical stages (e.g., Clinical Dementia Rating (CDR) ≥2) [2, 3]. Neuropsychiatric symptoms, including psychosis, increase caregiver burden considerably [4] and these patients often need additional attention leading to long-term care placement. The specific etiology for psychotic symptoms in AD has yet to be elucidated and is likely complicated by the wide range of reported symptoms. Investigating biological and pathological brain changes in specific regions that are associated with a particular symptom will facilitate early diagnosis and better prognosis.

AD dementia is defined clinically as cognitive and functional impairment [5] and is confirmed histopathologically as amyloid-β plaques and neurofibrillary tau tangles in key brain regions at autopsy [6]. Tau pathology is more closely associated with neurodegeneration and clinical severity compared to amyloid pathology in AD [7]. Therefore, tau may be the pathogenic driver underlying the various neurodegeneration-related changes associated with psychotic symptoms. Post-mortem studies show that psychotic symptoms are associated with greater hyperphosphorylated tau and neurofibrillary tangle burden in the right frontal cortex [8].

In research, the AD continuum is defined biologically as the presence of amyloid-β and phosphorylated tau or their aggregates that follows a stereotyped spatial and temporal progression [9, 10], even prior to downstream neurodegeneration and cognitive impairment [7, 11]. *In vivo* neuroimaging provides critical spatial and temporal information about AD pathology in relation to symptomatology development. Further, neuroimaging provides information across the entire brain, compared to selective *a priori* slices at autopsy. Previous work has demonstrated hypometabolism in the right lateral frontal cortex in AD patients with delusions [12] and atrophy in the insula, hippocampus, and amygdala in AD patients with hallucinations [13], but the cause of those neurodegeneration-related changes still remains unclear. Only one study has investigated living AD patients with tau PET as a potential pathological driver of psychotic symptoms; however the investigators did not correct for clinical severity, and in that study AD patients with psychotic symptoms had worse CDR scores and greater tau burden in Braak stage regions than AD patients without psychotic symptoms. Therefore, it remains unknown if psychotic symptoms in AD is more related to overall disease progression or tau pathology itself [14].

In this study we assessed differences in tau burden between patients with psychotic symptoms (+P) and those without psychotic symptoms (-P) using PET scans from the Alzheimer’s Disease Neuroimaging Initiative (ADNI) database. We utilized a matching approach to find participants with similar clinical severity to evaluate the specific association of tau deposition with psychotic symptoms, above and beyond the tau deposition across Braak stage regions characteristic of AD progression. We hypothesized that composite Braak regions associated with AD will not differ between CDR matched groups with and without psychosis, but that individual regions, particularly limbic and frontal regions, will have increased tau PET burden.

## Methods

### Participants

Data used in the preparation of this article were obtained from the Alzheimer’s Disease Neuroimaging Initiative (ADNI) database (adni.loni.usc.edu). The ADNI was launched in 2003 as a public-private partnership, led by Principal Investigator Michael W. Weiner, MD. The primary goal of ADNI has been to test whether serial magnetic resonance imaging (MRI), positron emission tomography (PET), other biological markers, and clinical and neuropsychological assessment can be combined to measure the progression of mild cognitive impairment (MCI) and early Alzheimer’s disease (AD). For up-to-date information, see www.adni-info.org.

We included participants who underwent tau PET imaging, amyloid PET imaging, structural T1 MR imaging, and CDR evaluation. As per ADNI inclusion criteria, participants were excluded if they had a long history of ongoing psychotic symptoms associated with other psychiatric disorders [15]. We identified 26 participants (+P) who endorsed psychotic symptoms (i.e., delusions and/or hallucinations), based on the NeuroPsychiatric Inventory (NPI) [16] or the NeuroPsychiatric Inventory Questionnaire (NPI-Q) [17] depending on their ADNI visit. We used participant and/or co-participant responses to NPI/NPI-Q questions A and B, which ask for a binary endorsement of delusions and hallucinations, respectively. If an endorsement is made, severity is also assessed; however due to the truncated range in our sample, severity was not assessed as part of this analysis. We then matched participants who did not endorse either psychotic symptom (n=26; -P) manually by creating a demographic profile for each +P participant and matched each with a -P participant based on CDR, age, gender, race and ethnicity, eliminating potential matches sequentially at each demographic category. For the analysis, we used neuroimaging data and CDR evaluations that were nearest to the NPI/NPIQ data.

### MRI acquisition

Structural T1 MR imaging was conducted under the ADNI protocol (3 Tesla; magnetization prepared rapid gradient echo (MPRAGE) sequence: repetition time (TR)1=12300 ms, echo time (TE)1=1minimum full echo, inversion time (TI)1=1900 ms, field of view (FOV)1=12081×12401×12561mm, voxel resolution = 11×111×111mm). Pmod (version 4.2; Pmod Technologies LLC, Switzerland) anatomical segmentation (cortical Hammers-N30R83-1MM and cerebellar AAL-1MM atlas) of the T1 scan was used for tau PET quantification in volume-weighted bilateral regions of interest (ROIs), including hippocampus, amygdala, frontal lobe, temporal lobe, and parietal lobe. Composite Braak I/II (Hippocampus and parahippocampus), Braak III/IV (fusiform gyrus, lingual gyrus and amygdala), and Braak V/VI (Frontal, Superior Temporal and Parietal Lobes) ROIs were modified from previously published Freesurfer-based segmentations to better reflect PMOD-based segmentations [18, 19].

### PET acquisition

[18F]AV1451 tau PET imaging was conducted under the ADNI protocol (370 MBq (10 mCi); 30-minute dynamic scan, comprised of six 5-minute frames acquired 75 minutes post-injection). Using Pmod (version 4.2, Pmod Technologies LLC, Switzerland), we calculated standard uptake value ratio (SUVR; 75-115 min post-injection; inferior cerebellar gray matter reference region) on the voxel-level.

[18F]Florbetaben and [18F]Florbetapir ([18F]AV45) amyloid PET imaging was conducted under the ADNI protocol ([18F]Florbetaben: 300 MBq (8.1 mCi); four 5-minute frames acquired 90 minutes post-injection; [18F]Florbetapir: 370 MBq (10.0 mCi); four 5-minute frames acquired 50 minutes post-injection). Amyloid PET SUVR was calculated within a global composite ROI including Thal phase regions and was harmonized across amyloid PET tracers into the Centiloid scale as previously described [20].

### Statistical analysis

We assessed our hypothesis that tau burden would be elevated in the +P group compared to CDR-matched -P group through a series of analyses. First, we performed paired t-tests on the ROI level, adjusting for global amyloid burden. Second, we performed a paired t-test on the voxel-level, adjusting for global amyloid burden. Adjusting for amyloid burden further removed variance in tau burden due to AD in a similar manner as CDR matching +P and -P groups, but avoids limiting the sample size via exact matching and unwanted variance from inexact matching. Performing a voxelwise analysis (unadjusted p-value<0.01) allowed the identification of additional regions where tau burden may be elevated in the +P group compared to the CDR matched -P group in (a) non-Braak regions, (b) Braak regions with a unilateral pattern that may be attenuated in bilateral ROI analyses, and (c) individual Braak regions that may be attenuated in composite Braak ROI analyses.

Because +P participants could have endorsed psychotic symptoms on NPI/NPI-Q before, after, or concurrent with their tau scan (i.e. within a year of tau PET), and could have endorsed only delusions, only hallucinations or both delusions and hallucinations, we ran exploratory analyses breaking down the timing relative to tau imaging and the type of psychotic symptom. Therefore, there were five groups in the exploratory analyses – no psychotic symptoms (-P), psychotic symptoms endorsed after tau PET (n=8), psychotic symptoms endorsed prior to tau PET (n=6), concurrent delusions (n=9), and concurrent hallucinations (n=3). One participant endorsed concurrent delusions and concurrent hallucinations. They were included in the concurrent delusions group because that was the more common symptom overall, but sensitivity analyses including them in the concurrent hallucinations group did not change results. For a full breakdown of psychotic symptoms and timing relative to tau PET, see Supplemental tables 1 and 2. To evaluate subgroups, we ran an ANOVA on the ROI level, adjusting for global amyloid burden. Then, we descriptively present participant level scans as sample sizes in exploratory groups were too small for voxel-wise analyses. All ROI-level statistics were performed in R version 4.2.1 and all voxelwise statistics were performed in SPM12 (MATLAB).

**Table 1.** Demographic characteristics for the entire analytic sample and by individuals with no psychotic symptoms and individuals with psychotic symptoms.

|  | Total | Individuals with no<br>Psychotic Symptoms | Individuals with<br>Psychotic Symptoms |
| --- | --- | --- | --- |
| N | 52 | 26 | 26 |
| Age<br>Mean [SD] | 77 [7] | 77 [6] | 77 [7] |
| Female / Male | 25 / 27 | 12 / 14 | 13 / 13 |
| Education<br>Mean [SD] | 16 [3] | 15 [3] | 16 [3] |
| Race |  |  |  |
| % White | 92 | 92 | 92 |
| % Black/African<br>American | 4 | 4 | 4 |
| % Asian | 4 | 4 | 4 |
| Ethnicity |  |  |  |
| % Non-Hispanic | 94 | 92 | 96 |
| % Hispanic | 6 | 8 | 4 |
| Centiloids<br>Mean [SD] | 64.8 [46.1] | 60.5 [48.7] | 69.2 [43.9] |
| Global CDR |  |  |  |
| 0 | 6 | 3 | 3 |
| 0.5 | 22 | 11 | 11 |
| 1 | 16 | 8 | 8 |
| 2 | 6 | 3 | 3 |
| 3 | 2 | 1 | 1 |

**Table 2.** Tau burden (SUVR) group means and 95% confidence interval for individuals with no psychotic symptoms and individuals with psychotic symptoms, adjusted for amyloid burden, within different regions of interest (ROI). **Significant differences (p<0.01).

| ROI | Individuals with no Psychotic symptoms | Individuals with Psychotic Symptoms | T-statistic, p-value |
| --- | --- | --- | --- |
| Amygdala<br>*Mean [95% CI]* | 1.23<br>[1.03 – 1.44] | 1.48<br>[1.26 – 1.70] | T= 3.02, p= 0.006 ** |
| Hippocampus<br>*Mean [95% CI]* | 1.18<br>[1.05 – 1.32] | 1.31<br>[1.16 – 1.45] | T= 1.67, p= 0.1 |
| Braak I/II<br>*Mean [95% CI]* | 1.18<br>[1.02 – 1.33] | 1.31<br>[1.14 – 1.47] | T= 1.94, p=0.06 |
| Braak III/IV<br>*Mean [95% CI]* | 1.41<br>[1.19 – 1.63] | 1.54<br>[1.30 – 1.77] | T= 1.45, p=0.16 |
| Braak V/VI<br>*Mean [95% CI]* | 1.04<br>[0.89 – 1.19] | 1.08<br>[0.93 – 1.24] | T= 0.80, p= 0.43 |
| Frontal Lobe<br>*Mean [95% CI]* | 1.00<br>[0.86 – 1.14] | 1.07<br>[0.92 – 1.22] | T= 1.25, p=0.22 |
| Temporal Lobe<br>*Mean [95% CI]* | 1.19<br>[0.98 – 1.41] | 1.27<br>[1.04 – 1.50] | T= 0.81, p= 0.43 |
| Parietal Lobe<br>*Mean [95% CI]* | 1.09<br>[0.91 – 1.27] | 1.09<br>[0.90 – 1.28] | T= 0.03, p= 0.98 |

## Results

Fifty-two participants from ADNI were evaluated in this study, with twenty-six reporting psychotic symptoms (+P) and twenty-six reporting no psychotic symptoms (-P), matched for CDR and demographics (Table 1). In this analytic sample, the average age was 77 years old, 52% were women, the mean education was 16 years, and the majority self-identified as Non-Hispanic White. Demographic variables were not different between groups by design. Similarly, global CDR was not different between groups, but ranged from CDR 0 to CDR 3. The majority of participants were CDR 0.5 (42%) and CDR 1 (31%). Global amyloid burden was non-significantly greater in the +P group compared to the -P group (Table 1).

After removing CDR and demographic effects through matching and amyloid effects through covariate adjustment, tau burden in the amygdala was significantly greater in the +P group compared to the -P group (Figure 1; Table 2). In complementary voxelwise analyses (Figure 2), clusters of elevated tau burden in the +P group compared to the -P group were evident bilaterally in the amygdala, thalamus, putamen, and unilaterally in the right hippocampus, right entorhinal cortex, and right frontal cortex. We also observed small clusters in the white matter, ventral pallidum, and superior cerebellum. Further breaking down the +P group by endorsed symptoms in relation to tau PET timing (Supplemental Table 1 and 2), tau burden in the amygdala was significantly greater in the concurrent delusions group compared to the -P group (Figure 3; Table 3). Participant level tau SUVR images (Figure 4) demonstrate variability in regionality (e.g., laterality) and magnitude that differs from typical Braak staging.

**Figure 1.**
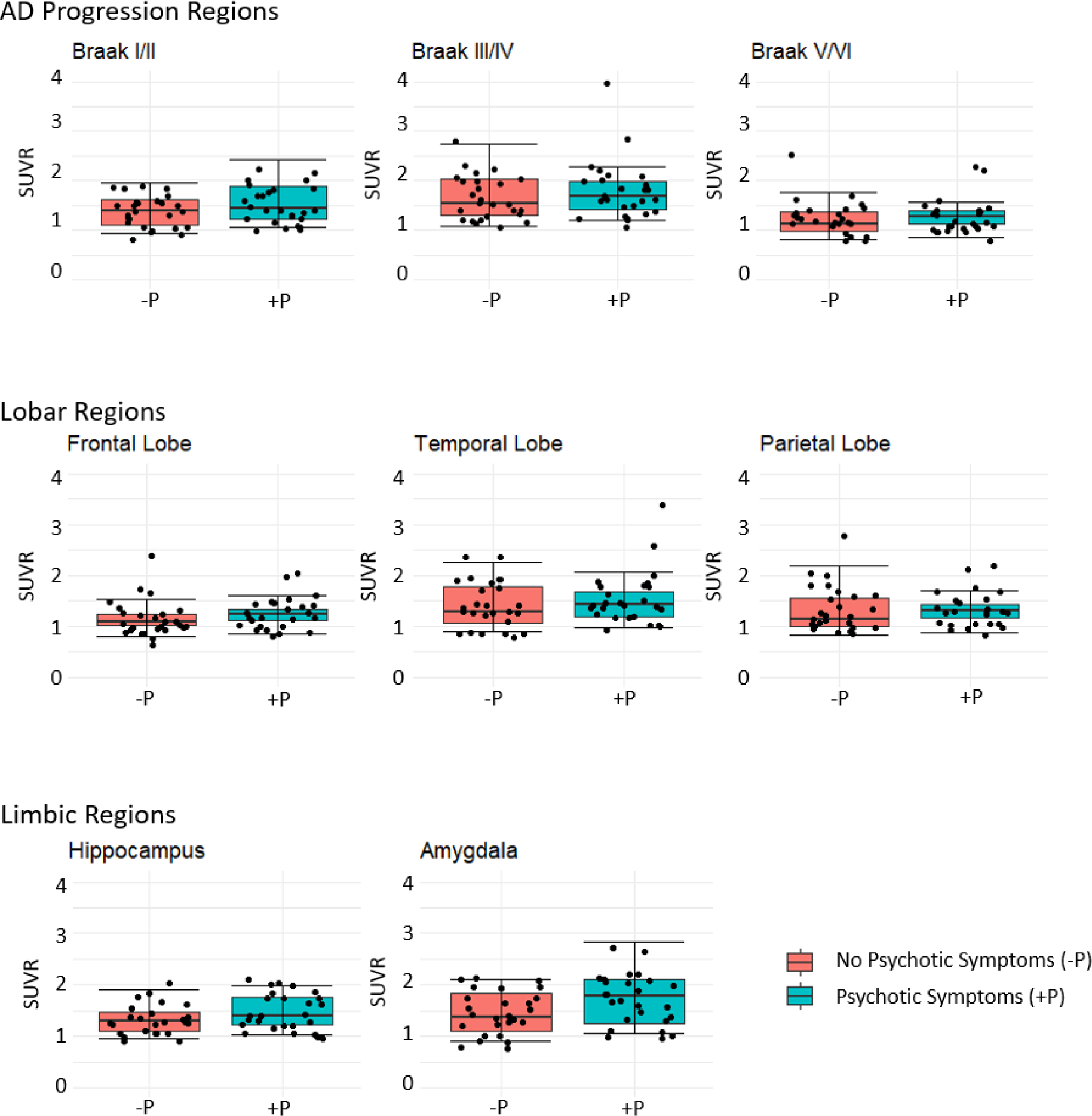
Distribution of tau burden (SUVR) between individuals with no psychotic symptoms (-P) and individuals with psychotic symptoms (+P), adjusted for amyloid burden.

**Figure 2:**
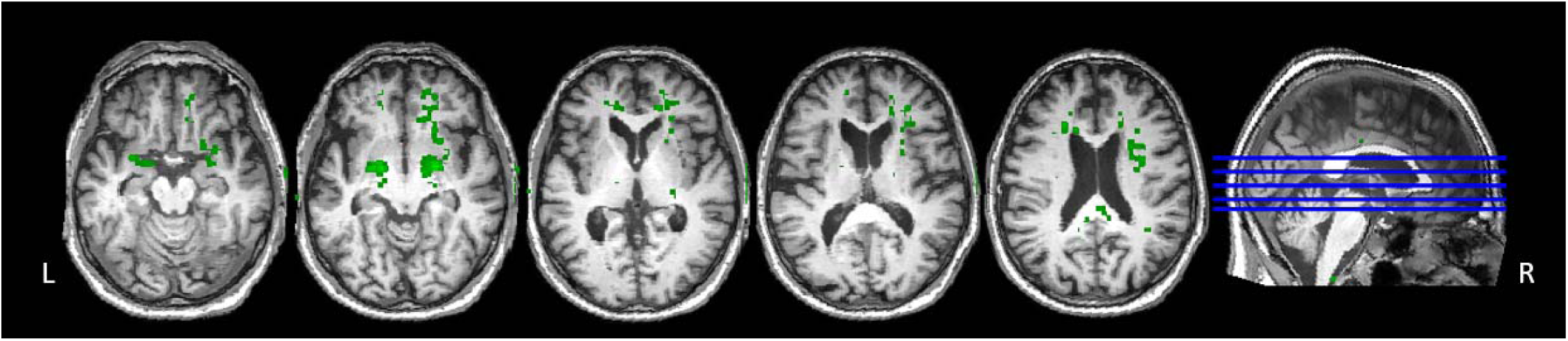
Voxelwise paired t-test between CDR and demographic matched individuals with psychotic symptoms (+P) and individuals without psychotic symptoms (-P), adjusted for amyloid burden. Voxels in green show areas of greater tau burden in the +P group compared to the -P group. No significant voxels found in the other direction (i.e., greater tau burden in the -P group compared to +P group). Significance was determined at p< 0.01 without multiple comparisons correction.

**Figure 3.**
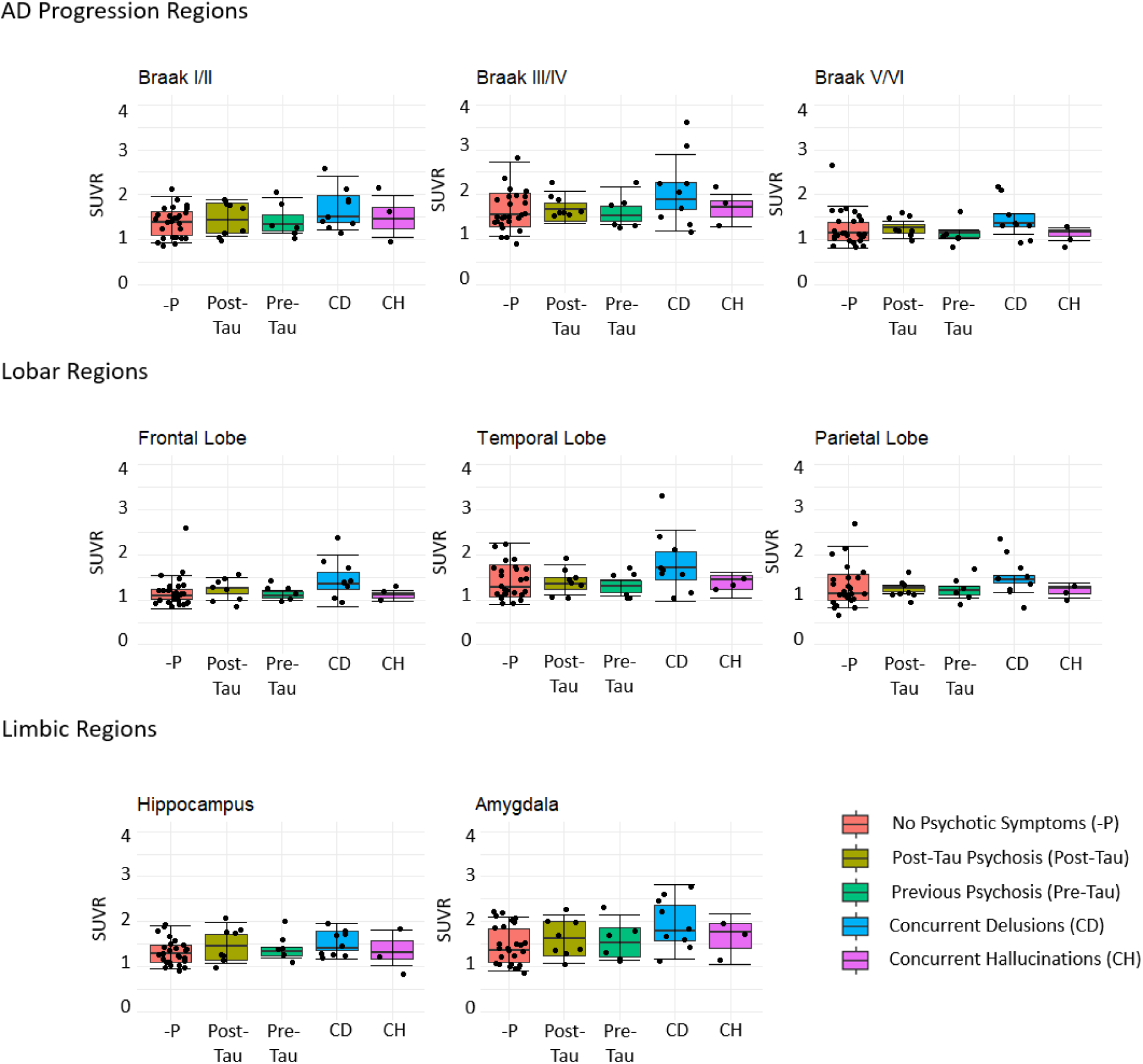
Distribution of tau burden (SUVR) between individuals with no psychotic symptoms (-P) and the Psychotic Symptom group categories: Post-Tau Psychotic Symptoms (Post-Tau), Previous Psychotic Symptoms (Pre-Tau), Concurrent Delusions (CD), and Concurrent Hallucinations (CH), adjusted for amyloid burden.

**Figure 4.**
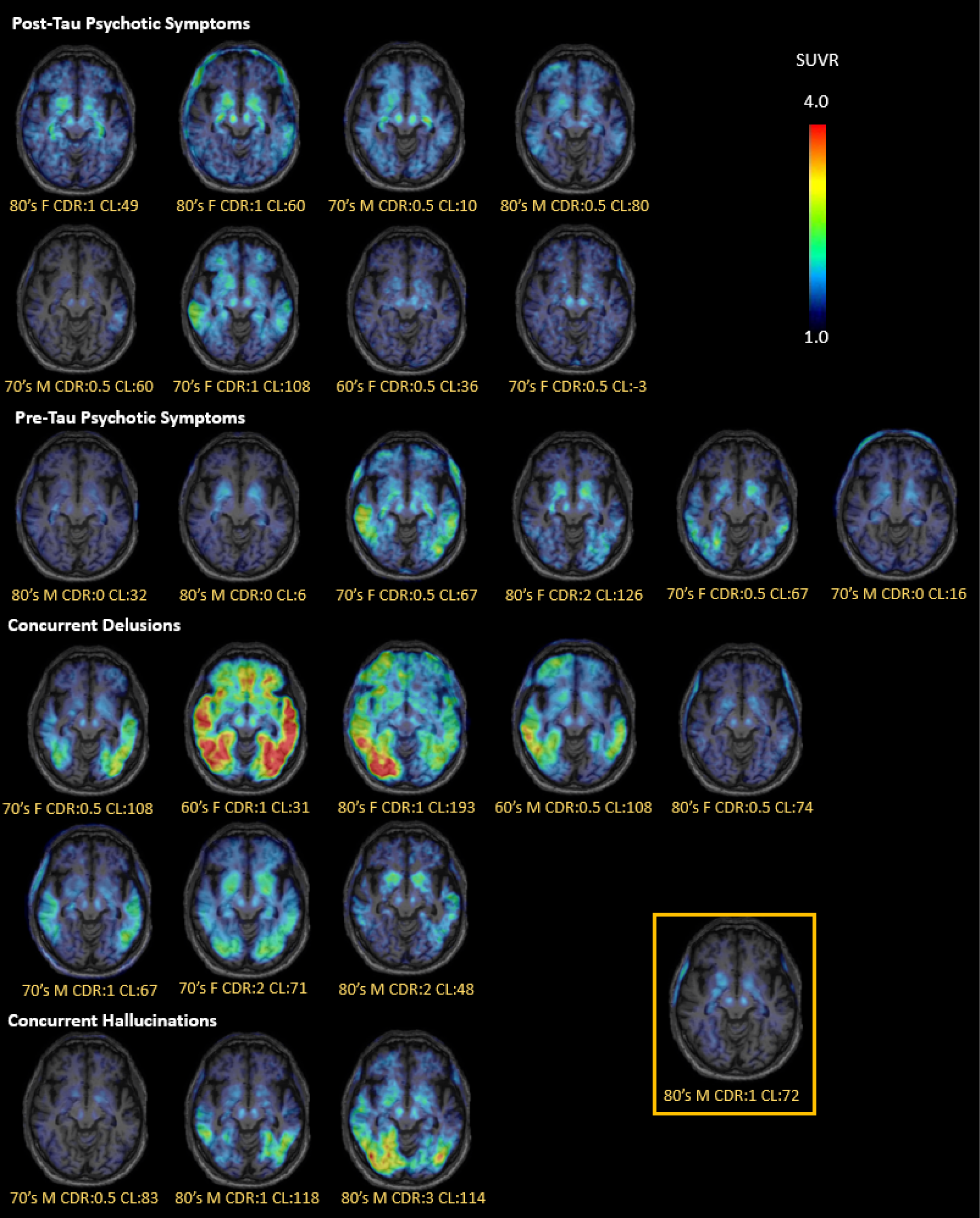
Representative slice from participant level tau SUVR scans in standardized space for exploratory psychotic symptoms subgroups. Age range is provided to prevent identification in addition to CDR and amyloid burden (Centiloid; CL). The yellow box indicates one participant endorsed both concurrent delusions and concurrent hallucinations, but was included in the concurrent delusions group for analyses.

**Table 3.** Tau burden (SUVR) group means and their 95% confidence interval for individuals with no psychotic symptoms, post-tau psychotic symptoms, pre-tau psychotic symptoms, concurrent delusions, and concurrent hallucinations, adjusted for amyloid burden, within different regions of interest (ROI). *Significant differences (p<0.05).

| ROI | Individuals with no Psychotic Symptoms | Individuals with Post-Tau Psychotic Symptoms | Individuals with Pre-Tau Psychotic Symptoms | Individuals with Concurrent Delusions | Individuals with Concurrent Hallucinations | F-statistic, p-value |
| --- | --- | --- | --- | --- | --- | --- |
| Amygdala<br>*Mean*<br>*95% CI* | 1.16<br>[0.95 -1.37] | 1.38<br>[1.08 -1.67] | 1.31<br>[0.97-1.64] | 1.54<br>[1.21-1.87] | 1.14<br>[0.63-1.65] | F= 2.9,<br>p= 0.03* |
| Hippocampus<br>*Mean*<br>*95% CI* | 1.18<br>[1.03-1.32] | 1.34<br>[1.14-1.54] | 1.27<br>[1.04-1.50] | 1.34<br>[1.11-1.56] | 1.12<br>[0.78-1.47] | F= 1.3<br>p= 0.27 |
| Braak I/II<br>*Mean*<br>*95% CI* | 1.14<br>[0.98-1.30] | 1.27<br>[1.05-1.49] | 1.21<br>[0.96-1.46] | 1.36<br>[1.11-1.60] | 1.07<br>[0.69-1.44] | F= 2.1<br>p= 0.09 |
| Braak III/IV<br>*Mean*<br>*95% CI* | 1.37<br>[1.13-1.60] | 1.42<br>[1.09-1.75] | 1.37<br>[1.00-1.75] | 1.72<br>[1.35-2.09] | 1.18<br>[0.61-1.74] | F= 2.3<br>p= 0.08 |
| Braak V/VI<br>*Mean*<br>*95% CI* | 1.02<br>[0.86-1.17] | 1.07<br>[0.85-1.30] | 1.00<br>[0.74-1.25] | 1.18<br>[0.94-1.43] | 0.77<br>[0.39-1.15] | F= 1.6<br>p= 0.18 |
| Frontal Lobe<br>*Mean*<br>*95% CI* | 0.99<br>[0.84-1.14] | 1.08<br>[0.87-1.29] | 0.99<br>[0.75-1.23] | 1.83<br>[0.16-2.07] | 0.77<br>[0.40-1.13] | F= 2.0<br>p= 0.12 |
| Temporal Lobe<br>*Mean*<br>*95% CI* | 1.17<br>[0.95-1.39] | 1.16<br>[0.85-1.47] | 1.13<br>[0.77-1.48] | 1.50<br>[1.16-1.84] | 0.90<br>[0.37-1.44] | F= 2.5<br>p= 0.052 |
| Parietal Lobe<br>*Mean*<br>*95% CI* | 1.05<br>[0.86-1.24] | 1.07<br>[0.80-1.34] | 1.01<br>[0.70-1.32] | 1.15<br>[0.85-1.45] | 0.76<br>[0.29-1.22] | F= 0.91<br>p= 0.47 |

## Discussion

Neurofibrillary tau tangles, measured with tau PET, were elevated in the amygdala, but not typical composite Braak stage regions, above and beyond the level expected for age, amyloid burden, and AD clinical severity in older adults. In addition, tau burden in the thalamus, putamen, right hippocampus, right entorhinal cortex, and right frontal cortex may also contribute to the structural and functional changes commonly reported in those regions for individuals with psychotic symptoms [12, 21, 22]. As tau PET may become increasingly incorporated into the clinical workflow for the early diagnosis of patients with AD, it will be critical to understand how tau pathology may contribute to symptom development and progression beyond cognitive and functional impairment to reduce patient and caregiver burden. High tau PET burden could signal an increased risk of psychosis, which may necessitate a more proactive approach to symptom management through medication and caregiver education.

Our results add to the growing body of work that aims to elucidate the neurobiological correlates of psychotic symptoms. In relation to psychosis [23, 24], most studies investigate structural biomarkers, including gray matter atrophy in neurodegenerative disease [13], white matter connectivity in schizophrenia [25], and neuronal metabolism in AD [12]. These neurodegeneration-related changes are often localized to frontal and limbic regions, but there is still considerable variability in relation to type of psychotic symptom and symptom severity in AD [26]. More recently, neuroinflammation has been investigated with symptom-specific findings in several neurodegenerative diseases [27].

While prior studies have investigated markers of neurodegeneration or inflammation, we sought to explore neurofibrillary tau burden as a potential pathological driver of psychotic symptoms in AD. We were interested in tau pathology rather than amyloid pathology because tau burden is more closely associated with neurodegeneration and symptomatic progression than amyloid burden. In this sample, there was no difference in amyloid burden in those with psychotic symptoms compared to those without. Previous work has identified elevated tau PET burden across Braak stage regions without correction for disease severity [14]. By adjusting for AD progression (i.e., CDR and amyloid burden) to understand psychosis-specific tau burden that is independent of AD clinical severity, we found evidence of elevated tau in the amygdala, but no evidence of elevated tau across composite Braak stage regions.

While many studies have recapitulated Braak staging with *in vivo* tau PET imaging in AD [7], there is still heterogeneity. Tau pathology of the AD type (i.e., 3R/4R isoform, paired helical filaments, neurofibrillary tangles) is found in older individuals without cognitive impairment [28–30], individuals with primary age related tauopathy [31], individuals with traumatic brain injury [32], specific mutations of frontotemporal dementia [33, 34], and dementia with Lewy bodies (DLB) [35, 36]. The magnitude and spatial distribution of AD-type tau pathology is relevant for different symptom profiles, even outside of the AD continuum, which motivated the complementary voxelwise analyses. Clusters of elevated tau in those with psychotic symptoms compared to those without were observed in the amygdala, thalamus, putamen, right hippocampus, right entorhinal cortex, and right frontal cortex. Smaller clusters in the superior cerebellum and white matter may be noise-related at this significance level and sample size, but other studies have demonstrated brain changes in the cerebellum [37] and white matter [25, 38] in relation to psychosis in non-AD patients.

The amygdala is often implicated in neuroimaging studies of psychosis because of its role in moderating the stress response to an emotionally salient environmental cue (e.g., delusions, hallucinations) [39–41]. Limbic regions have neuronal projections through the thalamus and basal ganglia to cortical regions, including the fronto-temporal lobes that are also involved in the salience network [42]. Elevated tau burden in these regions may directly contribute to psychotic symptoms and faster clinical progression; alternatively, psychosis can disrupt health seeking behaviors and exacerbate medical comorbidities in AD patients, which could in turn with psychosis could increase risk of faster clinical progression.

Limitations of the study include the relatively small sample size, cross-sectional analyses, potential for common comorbidities driving psychotic symptoms, and lack of information about psychotic symptom type, onset, duration, severity, and medication use. Despite the small cross-sectional sample, our results converge with other studies that identify neurobiological changes in the amygdala [43] and hippocampus [41], thalamus [44], putamen [45], and fronto-temporal lobes [12] as well as increasing dependence and faster cognitive decline in individuals with concurrent delusions compared to hallucinations [46]. However, larger longitudinal tau PET studies of delusions and hallucinations are warranted to make definitive conclusions. Psychotic symptoms often develop later in the AD continuum (e.g., CDR 2), but were present from CDR 0 to 3 in this ADNI sample. Including CDR 0 limits our conclusions related to psychotic symptoms in AD, but supports our conclusions related to tau pathology as an etiologic driver of psychotic symptoms in aging and disease. In ADNI, there was no information related to DLB, which is commonly characterized with psychotic symptoms, in order to understand the level of comorbidity. We cannot rule out DLB as the driver of psychotic symptoms in these individuals, but the ADNI recruitment criteria may minimize this possibility by excluding any significant neurological disease other than AD (e.g., Parkinson’s disease, multi-infarct dementia, Huntington’s disease, normal pressure hydrocephalus, brain tumor, progressive supranuclear palsy, seizure disorder, subdural hematoma, multiple sclerosis, history of significant head trauma followed by persistent neurological deficits, known structural brain abnormalities) as well as any significant systemic illness or unstable medical condition. Even if DLB was present in some of these individuals, the interaction between Lewy bodies and neurofibrillary tau tangles [47], particularly in the amygdala [48], suggests that tau PET may be a meaningful biomarker for psychosis. Larger longitudinal tau PET studies should explicitly consider the symptom type, onset, duration, and severity, as well as concomitant medication use.

Overall, a pathological biomarker for psychotic symptoms in AD could aid in early diagnosis, inform therapeutic agents in clinical trials, and complement existing neurobiological biomarkers (e.g., neuroinflammation, neuronal metabolism, white matter connectivity, gray matter atrophy). Elevated tau burden, above and beyond what is expected with age, amyloid burden, and clinical AD severity, in key brain regions provides insight into the various neurodegenerative and neuroinflammatory profiles reported in individuals with psychotic symptoms. There may not be one single cause of psychosis, but many contributing factors, and neurofibrillary tau tangles in limbic regions (e.g., amygdala) involved in emotional processing may represent one pathway to psychotic symptom onset.

## Data Availability

All data produced in the present study are available upon reasonable request to the authors and by request made to ADNI.

https://adni.loni.usc.edu/data-samples/access-data/

## Acknowledgement

Data collection and sharing for this project was funded by the Alzheimer’s Disease Neuroimaging Initiative (ADNI) (National Institutes of Health Grant U01 AG024904) and DOD ADNI (Department of Defense award number W81XWH-12-2-0012). ADNI is funded by the National Institute on Aging, the National Institute of Biomedical Imaging and Bioengineering, and through generous contributions from the following: AbbVie, Alzheimer’s Association; Alzheimer’s Drug Discovery Foundation; Araclon Biotech; BioClinica, Inc.; Biogen; Bristol-Myers Squibb Company; CereSpir, Inc.; Cogstate; Eisai Inc.; Elan Pharmaceuticals, Inc.; Eli Lilly and Company; EuroImmun; F. Hoffmann-La Roche Ltd and its affiliated company Genentech, Inc.; Fujirebio; GE Healthcare; IXICO Ltd.;Janssen Alzheimer Immunotherapy Research & Development, LLC.; Johnson & Johnson Pharmaceutical Research & Development LLC.; Lumosity; Lundbeck; Merck & Co., Inc.;Meso Scale Diagnostics, LLC.; NeuroRx Research; Neurotrack Technologies; Novartis Pharmaceuticals Corporation; Pfizer Inc.; Piramal Imaging; Servier; Takeda Pharmaceutical Company; and Transition Therapeutics. The Canadian Institutes of Health Research is providing funds to support ADNI clinical sites in Canada. Private sector contributions are facilitated by the Foundation for the National Institutes of Health (www.fnih.org). The grantee organization is the Northern California Institute for Research and Education, and the study is coordinated by the Alzheimer’s Therapeutic Research Institute at the University of Southern California. ADNI data are disseminated by the Laboratory for Neuro Imaging at the University of Southern California.

Daniel Talmasov, MD was supported by a NIMH T32 research fellowship in late-life neuropsychiatric disorders: NIH/NIMH Project #5T32MH020004-22.

This work was additionally supported by R00AG065506

## References

1. Cummings, J., et al., Criteria for psychosis in major and mild neurocognitive disorders: International Psychogeriatric Association (IPA) consensus clinical and research definition. 2020. 28(12): p. 1256–1269.

2. Hirono, N., et al., Factors associated with psychotic symptoms in Alzheimer’s disease. 1998. 64(5): p. 648–652.

3. Ismail, Z., et al., Psychosis in Alzheimer disease—mechanisms, genetics and therapeutic opportunities. 2022. 18(3): p. 131–144.

4. Kaufer, D.I., et al., Assessing the impact of neuropsychiatric symptoms in Alzheimer’s disease: the Neuropsychiatric Inventory Caregiver Distress Scale. 1998. 46(2): p. 210–215.

5. McKhann, G.M., et al., The diagnosis of dementia due to Alzheimer’s disease: Recommendations from the National Institute on Aging-Alzheimer’s Association workgroups on diagnostic guidelines for Alzheimer’s disease. 2011. 7(3): p. 263–269.

6. Montine, T.J., et al., National Institute on Aging–Alzheimer’s Association guidelines for the neuropathologic assessment of Alzheimer’s disease: a practical approach. 2012. 123: p. 1–11.

7. Jack Jr, C.R., et al., NIA-AA research framework: toward a biological definition of Alzheimer’s disease. 2018. 14(4): p. 535–562.

8. Koppel, J., et al., Psychotic Alzheimer’s disease is associated with gender-specific tau phosphorylation abnormalities. 2014. 35(9): p. 2021–2028.

9. Thal, D.R., et al., Phases of Aβ-deposition in the human brain and its relevance for the development of AD. 2002. 58(12): p. 1791–1800.

10. Braak, H. and E.J.A.n. Braak, Neuropathological stageing of Alzheimer-related changes. 1991. 82(4): p. 239–259.

11. Jack Jr, C.R., et al., Introduction to revised criteria for the diagnosis of Alzheimer’s disease: National Institute on Aging and the Alzheimer Association Workgroups. 2011. 7(3): p. 257.

12. Sultzer, D.L., et al., Neurobiology of delusions, memory, and insight in Alzheimer disease. 2014. 22(11): p. 1346–1355.

13. Blanc, F., et al., Right anterior insula: core region of hallucinations in cognitive neurodegenerative diseases. 2014. 9(12): p. e114774.

14. Gomar, J.J., et al., Increased retention of tau PET ligand [18F]-AV1451 in Alzheimer’s Disease Psychosis. 2022. 12(1): p. 82.

15. Mueller, S.G., et al., The Alzheimer’s disease neuroimaging initiative. 2005. 15(4): p. 869–877.

16. Cummings, J.J.J.o.g.p. and neurology, The neuropsychiatric inventory: development and applications. 2020. 33(2): p. 73–84.

17. Kaufer, D.I., et al., Validation of the NPI-Q, a brief clinical form of the Neuropsychiatric Inventory. 2000. 12(2): p. 233–239.

18. Kreisl, W.C., et al., Patterns of tau pathology identified with 18F-MK-6240 PET imaging. 2022. 18(2): p. 272–282.

19. Schöll, M., et al., PET imaging of tau deposition in the aging human brain. 2016. 89(5): p. 971–982.

20. Royse, S.K., et al., Validation of amyloid PET positivity thresholds in centiloids: a multisite PET study approach. 2021. 13(1): p. 99.

21. Koppel, J., et al., Psychosis in Alzheimer’s Disease is Associated with Frontal Metabolic Impairment and Accelerated Decline in Working Memory: Findings from the Alzheimer’s Disease Neuroimaging Initiative. 2014. 22(7): p. 698–707.

22. Qian, W., et al., Gray matter changes associated with the development of delusions in Alzheimer disease. 2019. 27(5): p. 490–498.

23. Corlett, P.R., et al., Toward a neurobiology of delusions. 2010. 92(3): p. 345–369.

24. Allen, P., et al., The hallucinating brain: a review of structural and functional neuroimaging studies of hallucinations. 2008. 32(1): p. 175–191.

25. Davis, K.L., et al., White matter changes in schizophrenia: evidence for myelin-related dysfunction. 2003. 60(5): p. 443–456.

26. Lyketsos, C.G., et al., Neuropsychiatric symptoms in Alzheimer’s disease. 2011, Elsevier. p. 532–539.

27. Dunn, G.A., et al., Neuroinflammation in psychiatric disorders: An introductory primer. 2020. 196: p. 172981.

28. Ossenkoppele, R., et al., Amyloid and tau PET-positive cognitively unimpaired individuals are at high risk for future cognitive decline. 2022. 28(11): p. 2381–2387.

29. Chen, X., et al., Regional tau effects on prospective cognitive change in cognitively normal older adults. 2021. 41(2): p. 366–375.

30. Ziontz, J., et al., Tau pathology in cognitively normal older adults. 2019. 11: p. 637–645.

31. Bell, W.R., et al., Neuropathologic, genetic, and longitudinal cognitive profiles in primary age-related tauopathy (PART) and Alzheimer’s disease. 2019. 15(1): p. 8–16.

32. Ayubcha, C., et al., A critical review of radiotracers in the positron emission tomography imaging of traumatic brain injury: FDG, tau, and amyloid imaging in mild traumatic brain injury and chronic traumatic encephalopathy. 2021. 48: p. 623–641.

33. Tsai, R.M., et al., 18 F-flortaucipir (AV-1451) tau PET in frontotemporal dementia syndromes. 2019. 11: p. 1–18.

34. Levy, J.P., et al., 18F-MK-6240 tau-PET in genetic frontotemporal dementia. 2022. 145(5): p. 1763–1772.

35. Gomperts, S.N., et al., Tau positron emission tomographic imaging in the Lewy body diseases. 2016. 73(11): p. 1334–1341.

36. Chin, K.S., et al., Prevalence and clinical associations of tau in Lewy body dementias: A systematic review and meta-analysis. 2020. 80: p. 184–193.

37. Krämer, J., et al., Abnormal cerebellar volume in somatic vs. non-somatic delusional disorders. 2020. 7(1): p. 1–8.

38. Takahata, K., et al., PET-detectable tau pathology correlates with long-term neuropsychiatric outcomes in patients with traumatic brain injury. 2019. 142(10): p. 3265–3279.

39. Fudge, J.L., et al., Considering the role of the amygdala in psychotic illness: a clinicopathological correlation. 1998. 10(4): p. 383–394.

40. Delavari, F., et al., Amygdala subdivisions exhibit aberrant whole-brain functional connectivity in relation to stress intolerance and psychotic symptoms in 22q11. 2DS. 2023. 13(1): p. 145.

41. Provenzano, F.A., et al., Hippocampal pathology in clinical high-risk patients and the onset of schizophrenia. 2020. 87(3): p. 234–242.

42. Solé-Padullés, C., et al., Intrinsic functional connectivity of fronto-temporal networks in adolescents with early psychosis. 2017. 26: p. 669–679.

43. Jaramillo-Jimenez, A., et al., Association between amygdala volume and trajectories of neuropsychiatric symptoms in Alzheimer’s disease and dementia with lewy bodies. 2021. 12: p. 679984.

44. Marshall, G.A., et al., Positron emission tomography metabolic correlates of apathy in Alzheimer disease. 2007. 64(7): p. 1015–1020.

45. Luo, X., et al., Putamen gray matter volumes in neuropsychiatric and neurodegenerative disorders. 2019. 3(1).

46. Kociolek, A.J., et al., Neuropsychiatric Symptoms and Trajectories of Dependence and Cognition in a Sample of Community-dwelling Older Adults with Dementia. 2023. 20(6): p. 409–419.

47. Gerson, J.E., et al., Tau oligomers mediate α-synuclein toxicity and can be targeted by immunotherapy. 2018. 13(1): p. 1–14.

48. Colom-Cadena, M., et al., Regional overlap of pathologies in Lewy body disorders. 2017. 76(3): p. 216–224.

