## Supplemental Tables 1 and 2 for "Psychotic symptoms are associated with elevated tau PET signal in the amygdala independent of Alzheimer’s disease clinical severity and amyloid burden"

Affiliations:

*Data used in preparation of this article were obtained from the Alzheimer's Disease Neuroimaging Initiative (ADNI) database (adni.loni.usc.edu). As such, the investigators within the ADNI contributed to the design and implementation of ADNI and/or provided data but did not participate in analysis or writing of this report. A complete listing of ADNI investigators can be found at: <http://adni.loni.usc.edu/wpcontent/uploads/how_to_apply/ADNI_Acknowledgement_List.pdf>

**Supplemental Materials**

**Supplemental Table 1:**

|  | Total | Individuals with No Psychotic Symptoms | Psychotic Symptoms | | | | |
| --- | --- | --- | --- | --- | --- | --- | --- |
|  |  |  | All Individuals with Psychotic Symptoms | Individuals with Post-tau Psychotic Symptoms | Individuals with Pre-tau Psychotic Symptoms | Individuals with Concurrent Delusions | Individuals with Concurrent Hallucinations |
| N | 52 | 26 | 26 | 8 | 6 | 9 | 3 |
| Age  Mean [SD] | 77 [7] | 77 [7] | 77 [6] | 76 [7] | 79 [5] | 76 [7] | 83 [3] |
| Female / Male | 27 / 25 | 14 / 12 | 13 / 13 | 5 / 3 | 3 / 3 | 5 / 4 | 0 / 3 |
| Education  Mean [SD] | 16 [3] | 15 [3] | 16 [3] | 15 [3] | 16 [3] | 16 [2] | 18 [2] |
| Race | | | | | | | |
| % White | 92 | 92 | 92 | 100 | 100 | 78 | 100 |
| % Black/African American | 4 | 4 | 4 | 0 | 0 | 11 | 0 |
| % Asian | 4 | 4 | 4 | 0 | 0 | 11 | 0 |
| Ethnicity | | | | | | | |
| % Non-Hispanic | 94 | 92 | 96 | 100 | 83 | 100 | 100 |
| % Hispanic | 6 | 8 | 4 | 0 | 17 | 0 | 0 |
| Centiloids  Mean [SD] | 64.8 [46.1] | 60.5  [48.7] | 69.2 [43.9] | 49.9 [36.1] | 52.5  [44.2] | 85.6  [47.1] | 105  [19] |
| Global CDR | | | | | | | |
| 0 | 6 | 3 | 3 | 0 | 3 | 0 | 0 |
| 0.5 | 22 | 11 | 11 | 5 | 2 | 4 | 0 |
| 1 | 16 | 8 | 8 | 3 | 0 | 5 | 0 |
| 2 | 6 | 3 | 3 | 0 | 2 | 0 | 2 |
| 3 | 2 | 1 | 1 | 0 | 0 | 0 | 1 |
| CDR Sum of Boxes | 5 [4.5] | 4 [4.6] | 5.5 [4.3] | 4 [1.8] | 3 [4.7] | 7.5 [3.9] | 8.5 [7] |
| MMSE | 23 [6] | 24 [6] | 23[6] | 25 [2] | 26 [6] | 19 [5] | 23 [7] |

Demographic characteristics for the entire analytic sample, by individuals with no Psychotic Symptoms, and by the Psychotic Symptom subgroups. CDR Sum of Boxes and MMSE, as measures of general cognitive function, were not significantly different across groups.

**Supplemental Table 2:**

| **ID** | **Psychotic Symptoms Description** | **Severity from 0-3**  **(Delusions, Hallucinations)** | **Number of Days from Tau scan to endorsement of psychotic symptoms** | **Category Designation** | **Global CDR** | **CDR SoB** | **MMSE** |
| --- | --- | --- | --- | --- | --- | --- | --- |
| 1 | Concurrent Delusions | 1, 0 | 27 | Concurrent Delusions | 0.5 | 4.5 | 24 |
| 2 | Concurrent Delusions | 1, 0 | 7 | Concurrent Delusions | 2 | 14 | 16 |
| 3 | Concurrent Delusions | 1, 0 | 80 | Concurrent Delusions | 1 | 7 | 19 |
| 4 | Concurrent Delusions | 2, 0 | 15 | Concurrent Delusions | 1 | 8 | 25 |
| 5 | Concurrent Delusions | 2, 0 | 12 | Concurrent Delusions | 2 | 14 | 9 |
| 6 | Concurrent Hallucinations and Delusions | 2, 2 | -313 | Concurrent Delusions | 1 | 5 | 16 |
| 7 | Concurrent Delusions | 1, 0 | 2 | Concurrent Delusions | 0.5 | 4.5 | 25 |
| 8 | Concurrent Delusions | 1, 0 | -8 | Concurrent Delusions | 0.5 | 4 | 19 |
| 9 | Concurrent Delusions | 3, 0 | 20 | Concurrent Delusions | 1 | 6.5 | 15 |
| 10 | Concurrent Hallucinations | 0, 1 | 24 | Concurrent Hallucinations | 1 | 8 | 22 |
| 11 | Concurrent Hallucinations | 0, 1 | 3 | Concurrent Hallucinations | 3 | 16 | 16 |
| 12 | Concurrent Hallucinations | 0, 1 | 42 | Concurrent Hallucinations | 0.5 | 2 | 30 |
| 13 | Post-Tau Hallucinations | 0, 1 | -1090 | Post-Tau Psychotic Symptoms | 1 | 6 | 24 |
| 14 | Post-Tau Delusions | 1, 0 | -749 | Post-Tau Psychotic Symptoms | 1 | 5.5 | 23 |
| 15 | Post-Tau Hallucinations and Delusions | 3, 1 | -1663 | Post-Tau Psychotic Symptoms | 0.5 | 1.5 | 26 |
| 16 | Post-Tau Delusions | 1, 0 | -1072 | Post-Tau Psychotic Symptoms | 0.5 | 2 | 24 |
| 17 | Post-Tau Delusions | 1, 0 | -1371 | Post-Tau Psychotic Symptoms | 0.5 | 3.5 | 28 |
| 18 | Post-Tau Hallucinations | 0, 1 | -728 | Post-Tau Psychotic Symptoms | 0.5 | 2.5 | 23 |
| 19 | Post-Tau Delusions | 1, 0 | -714 | Post-Tau Psychotic Symptoms | 1 | 6 | 25 |
| 20 | Post-Tau Hallucinations | 0, 1 | -383 | Post-Tau Psychotic Symptoms | 0.5 | 4.5 | 24 |
| 21 | Pre-Tau Hallucinations | 0, 1 | 4228 | Pre-Tau Psychotic Symptoms | 0 | 0 | 29 |
| 22 | Pre-Tau Delusions | 2, 0 | 3158 | Pre-Tau Psychotic Symptoms | 0 | 0 | 30 |
| 23 | Pre-Tau Hallucinations | 0, 1 | 1952 | Pre-Tau Psychotic Symptoms | 0.5 | 2.5 | 29 |
| 24 | Pre-Tau Delusions | 1, 0 | 1753 | Pre-Tau Psychotic Symptoms | 2 | 12 | 17 |
| 25 | Pre-Tau Delusions | 1, 0 | 1031 | Pre-Tau Psychotic Symptoms | 0 | 0 | 30 |
| 26 | Pre-Tau Delusions | 1, 0 | 375 | Pre-Tau Psychotic Symptoms | 0.5 | 4 | 18 |
| 27 | No Psychotic symptoms endorsed |  |  | No Psychotic Symptoms | 0 | 0 | 29 |
| 28 | No Psychotic symptoms endorsed |  |  | No Psychotic Symptoms | 3 | 15 | 19 |
| 29 | No Psychotic symptoms endorsed |  |  | No Psychotic Symptoms | 0 | 0 | 30 |
| 30 | No Psychotic symptoms endorsed |  |  | No Psychotic Symptoms | 1 | 4 | 23 |
| 31 | No Psychotic symptoms endorsed |  |  | No Psychotic Symptoms | 0.5 | 1.5 | 24 |
| 32 | No Psychotic symptoms endorsed |  |  | No Psychotic Symptoms | 2 | 12 | 10 |
| 33 | No Psychotic symptoms endorsed |  |  | No Psychotic Symptoms | 1 | 9 | 21 |
| 34 | No Psychotic symptoms endorsed |  |  | No Psychotic Symptoms | 1 | 4.5 | 21 |
| 35 | No Psychotic symptoms endorsed |  |  | No Psychotic Symptoms | 2 | 13 | 18 |
| 36 | No Psychotic symptoms endorsed |  |  | No Psychotic Symptoms | 0.5 | 0.5 | 28 |
| 37 | No Psychotic symptoms endorsed |  |  | No Psychotic Symptoms | 1 | 3.5 | 29 |
| 38 | No Psychotic symptoms endorsed |  |  | No Psychotic Symptoms | 0.5 | 0.5 | 28 |
| 39 | No Psychotic symptoms endorsed |  |  | No Psychotic Symptoms | 0 | 0 | 26 |
| 40 | No Psychotic symptoms endorsed |  |  | No Psychotic Symptoms | 1 | 8 | 20 |
| 41 | No Psychotic symptoms endorsed |  |  | No Psychotic Symptoms | 0.5 | 1.5 | 27 |
| 42 | No Psychotic symptoms endorsed |  |  | No Psychotic Symptoms | 1 | 6 | 18 |
| 43 | No Psychotic symptoms endorsed |  |  | No Psychotic Symptoms | 2 | 13 | 5 |
| 44 | No Psychotic symptoms endorsed |  |  | No Psychotic Symptoms | 0.5 | 0.5 | 28 |
| 45 | No Psychotic symptoms endorsed |  |  | No Psychotic Symptoms | 0.5 | 2.5 | 26 |
| 46 | No Psychotic symptoms endorsed |  |  | No Psychotic Symptoms | 0.5 | 1 | 29 |
| 47 | No Psychotic symptoms endorsed |  |  | No Psychotic Symptoms | 0.5 | 1.5 | 24 |
| 48 | No Psychotic symptoms endorsed |  |  | No Psychotic Symptoms | 0.5 | 1 | 28 |
| 49 | No Psychotic symptoms endorsed |  |  | No Psychotic Symptoms | 1 | 1 | 29 |
| 50 | No Psychotic symptoms endorsed |  |  | No Psychotic Symptoms | 1 | 3.5 | 21 |
| 51 | No Psychotic symptoms endorsed |  |  | No Psychotic Symptoms | 0.5 | 1.5 | 29 |
| 52 | No Psychotic symptoms endorsed |  |  | No Psychotic Symptoms | 0.5 | 1.5 | 28 |

Individual level data about psychotic symptoms, severity, timing relative to tau PET, subgroup designation, global CDR, CDR Sum of Boxes (as a measure of clinical severity), and MMSE (as a measure of general cognition). A duration ≤365 days between the endorsement of psychotic symptoms and the tau PET scan was considered to be concurrent.
